# Performance of Existing and Novel Surveillance Case Definitions for COVID-19 in the Community

**DOI:** 10.1101/2020.10.02.20195479

**Authors:** Hannah E. Reses, Mark Fajans, Scott H. Lee, Charles M. Heilig, Victoria T. Chu, Natalie J. Thornburg, Kim Christensen, Sanjib Bhattacharyya, Alicia Fry, Aron J. Hall, U.S. COVID-19 Household Investigation Team, Jacqueline E. Tate, Hannah L. Kirking, Scott A. Nabity

## Abstract

**Background:** Severe acute respiratory syndrome coronavirus 2 (SARS-CoV-2), the virus that causes coronavirus disease 2019 (COVID-19), presents with a broad range of symptoms. Existing COVID-19 case definitions were developed from early reports of severely ill, primarily hospitalized, patients. Symptom-based case definitions that guide public health surveillance and individual patient management in the community must be optimized for COVID-19 pandemic control.

**Methods:** We collected daily symptom diaries and performed RT-PCR on respiratory specimens over a 14-day period in 185 community members exposed to a household contact with COVID-19 in the Milwaukee, Wisconsin and Salt Lake City, Utah metropolitan areas. We interpreted the discriminatory performance (sensitivity, specificity, predictive values, F⍰1 score, Youden’s index, and prevalence estimation) of individual symptoms and common case definitions according to two principal surveillance applications (i.e., individual screening and case counting). We also constructed novel case definitions using an exhaustive search with over 73 million symptom combinations and calculated bias-corrected and accelerated bootstrap confidence intervals stratified by children versus adults.

**Findings:** Common COVID-19 case definitions generally showed high sensitivity (86⍰96%) but low positive predictive value (PPV) (36⍰49%; F⍰1 score 52⍰63) in this community cohort. The top performing novel symptom combinations included taste or smell dysfunction. They also improved the balance of sensitivity and PPV (F⍰1 score 78⍰80) and reduced the number of false positive symptom screens. Performance indicators were generally lower for children (<18 years of age).

**Interpretation:** Existing COVID-19 case definitions appropriately screened in community members with COVID-19. However, they led to many false positive symptom screens and poorly estimated community prevalence. Absent unlimited, timely testing capacity, more accurate case definitions may help focus public health resources. Novel symptom combinations incorporating taste or smell dysfunction as a primary component better balanced sensitivity and specificity. Case definitions tailored specifically for children versus adults should be further explored.

**Funding:** This research was wholly supported by the U.S. Centers for Disease Control and Prevention.

**Disclaimer:** The findings and conclusions in this report are those of the authors and do not necessarily represent the official position of the U.S. Centers for Disease Control and Prevention/the Agency for Toxic Substances and Disease Registry.

**Research in Context:** *Evidence before this study:* Coronavirus disease 2019 (COVID-19) incidence has accelerated globally over the last several months. As the full spectrum of clinical presentations has come into clearer focus, symptom-based clinical screening and case surveillance has also evolved. Preliminary understanding of the clinical manifestation of COVID-19 was driven primarily by descriptions of hospitalized patients, as early testing algorithms prioritized more severely ill persons with classic lower respiratory symptoms and fever. Since then, more data from ambulatory settings have emerged. We searched PubMed from 1 December 2019 to 21 August 2020 for studies that assessed the diagnostic performance of case surveillance definitions for COVID-19. We found no studies examining the discriminatory performance of case surveillance definitions among contacts with mild to moderate symptoms with documented exposure to persons with COVID-19. Nonetheless, we found nine highly relevant studies: seven original reports and two review articles. Five original studies evaluated individual, self-reported symptoms (two among healthcare workers in the United States, one among healthcare workers in the Netherlands, and one online survey for the general public in Somalia) and concluded that using dysfunction of taste or smell for routine COVID-19 screening likely had utility. The fifth study had a similar conclusion based on self-reported symptoms and laboratory results collected via smartphone from the general public in the United States and the United Kingdom. Another original study modeled the substantial effect that multiple revisions to the COVID-19 case definition had on the reported disease burden in the Chinese population. Lastly, an original study illustrated the shift in discriminatory performance of established influenza surveillance case definitions for influenza between adults and children. Age-specific differences in case definition performance may also apply to COVID-19. Two articles reviewed predictive algorithms to define outpatient COVID-19 illness and risk of hospitalization. The reviewed studies were limited in that they were either restricted to individual signs or symptoms, or they incorporated blood tests or imaging that required in-person access to medical care.

*Added value of this study:* The discriminatory performance of case surveillance definitions for COVID-19 is important for implementing effective epidemic mitigation strategies. Our study illustrates the performance of case definitions in community members with household exposure to severe acute respiratory syndrome coronavirus 2 (SARS-CoV-2) based solely on symptom profiles. Prior work overrepresented healthcare workers or otherwise studied non-representative populations, and they did not examine across the age spectrum. Our study also provides a novel framework for refining definitions. Using 15 symptoms associated with COVID-19 for all contacts regardless of disease status, we systematically evaluated the discriminatory performance of individual symptoms and previously defined case surveillance definitions across ages and according to two core surveillance applications: 1) screening non-hospitalized individuals to prioritize public health interventions, and 2) estimating the number of non-hospitalized persons with COVID-19 (i.e., community-based syndromic surveillance). We also constructed novel symptom combinations that effectively performed both functions and improved upon widely used case surveillance definitions that may help to target interventions in the absence of unlimited laboratory diagnostic capacity. Our analyses highlight the importance of ongoing re-evaluation of symptom-based surveillance definitions to suit the intended purpose and population under surveillance. Based on our results, which were derived from household members of all ages, case surveillance definition performance may improve if developed separately for adults and children.

*Implications of all the available evidence:* Case definitions for COVID-19 should be tailored to maximize the discriminatory performance dependent upon its intended use. Existing COVID-19 case definitions screened in most community members with COVID-19, but also yielded a high number of false positive results. When unlimited, timely diagnostic testing is not available symptom combinations with improved accuracy (i.e., more balanced sensitivity and specificity) may help focus resources, such as recommending self-isolation among community contacts.

## Background

Coronavirus disease 2019 (COVID-19) is caused by severe acute respiratory syndrome coronavirus 2 (SARS-CoV-2). The virus was first identified in a cluster of patients with atypical pneumonia in Wuhan, China, in December 2019.^1^ Since its emergence, the virus has spread globally, causing widespread infection and death. Following evidence of person-to-person transmission and a broader clinical spectrum of infections, case definitions for COVID-19 have been revised.^2^ In the initial weeks of the pandemic, COVID-19 was labeled a pneumonia of unknown etiology, and many who presented to medical care had classic pneumonia-like symptoms such as fever, cough, and dyspnea. An exceptional variety of symptoms has since been reported for COVID-19, ranging from none or mild indistinct symptoms to invasive neurological disease and fulminant respiratory failure.^3-7^ As is common in the early response phases to novel emerging pathogens, there is ongoing need to reassess and refine surveillance case definitions for COVID-19 based on new information. Changes to case definitions affect interpretation of surveillance data, as was demonstrated by substantially different prevalence estimates when China broadened the COVID-19 case definition early in its epidemic response.^2^

A few studies have demonstrated the predictive value of symptom profiles in healthcare workers^4,8,9^ and other populations potentially not necessarily representative of the general public.^5,10^ These studies are subject to other limitations, too. Some applied predictive models that included serum biomarkers and imaging.^11,12^ Obtaining this information may limit real-world capture of people with mild-to-moderate SARS-CoV-2 infection and may delay public health intervention. Further, few studies to date have examined symptom combinations exclusively. Respiratory pathogens routinely behave differently in children and adults, and this appears to be true for COVID-19 as well.^13^ For example, an assessment of ambulatory case surveillance definitions for influenza demonstrated lower sensitivity among children less than 5 years of age.^14^ Similar analyses across age strata are lacking for COVID-19. Reliable, age-stratified syndromic surveillance definitions would likely aid public health officials to scale up community contact tracing and develop protocols to safely operate various congregate venues, such as schools and workplaces, should unlimited, timely diagnostic testing be unavailable.

Dedicated symptom-based surveillance systems have been developed to track COVID-19 cases. These include the U.S. Council of State and Territorial Epidemiologists (CSTE) original (CSTE combination 1; released 5 April 2020) and revised (CSTE combination 2; released 7 August 2020) clinical criteria for reporting SARS-CoV-2 infection, and the original CDC COVID-19⍰like illness (CLI) definition (Table 1). Similarly, the Centers for Disease Control and Prevention (CDC) maintains a list of symptoms for priority SARS-CoV-2 testing. These COVID-19 case definitions and the priority testing symptom list are intended to capture as many persons with COVID-19 as possible with confirmatory testing. Finally, longstanding respiratory virus surveillance networks established to monitor influenza-like illnesses (ILI) and acute respiratory infection (ARI), which is used for community-based syndromic surveillance of respiratory syncytial virus by the World Health Organization (WHO), may be plausibly adaptable platforms for monitoring COVID-19. The performance characteristics and utility of these syndromic surveillance platforms for COVID-19 have not been well defined.^5^

**Table 1.** Existing COVID-19 case definitions, respiratory illness surveillance case definitions, and derived compound symptom combinations assessed for diagnostic performance in a community cohort of 185 individuals with household COVID-19 exposure in Utah and Wisconsin, United States, March–May 2020

| Case definition category | Case definition | Criteria |
| --- | --- | --- |
| Existing COVID-19 case definitions | U.S. Centers for Disease Control and Prevention (CDC) COVID-19-like illness (CLI)* | Fever, cough, or shortness of breath |
|  | CDC symptom list<sup>†</sup> | Fever or chills, cough, shortness of breath or difficulty breathing, fatigue, muscle or body aches, headache, new loss of taste or smell, sore throat, congestion or runny nose, nausea or vomiting, diarrhea |
|  | U.S. Council of State and Territorial Epidemiologists (CSTE) COVID-19 original clinical criteria (CSTE combination 1)<sup>‡</sup> | At least one of the following: cough, shortness of breath, or discomfort breathing, OR at least two of the following: fever, chills, rigors, myalgia, headache, sore throat, new olfactory disorder and taste disorders |
|  | CSTE COVID-19 revised clinical criteria (CSTE combination 2)<sup>§</sup> | At least one of the following: cough, shortness of breath, discomfort breathing, new olfactory disorder, new taste disorder, or at least two of the following: fever, chills, rigors, myalgia, headache, sore throat, nausea or vomiting, diarrhea, fatigue, congestion or runny nose |
| Existing respiratory illness surveillance case definitions | Influenza-like illness (ILI)<sup>□</sup> | Fever AND cough and/or sore throat |
|  | World Health Organization (WHO) acute respiratory infection (ARI) definition for community-based respiratory syncytial virus (RSV) surveillance<sup>¶</sup> | At least one of the following: shortness of breath or cough, sore throat, or coryza |
| Derived compound symptom combinations** | Combination 1 | Taste and/or smell dysfunction, OR one of the following: shortness of breath, myalgia, or fever or chills |
|  | Combination 2 | Taste and/or smell dysfunction or discomfort breathing, OR at least two of the following: shortness of breath, wheezing, or fever or chills |
|  | Combination 3 | Taste and/or smell dysfunction, OR at least two of the following: shortness of breath, wheezing, discomfort breathing, or fever or chills |
|  | Combination 4 | Taste and/or smell dysfunction, OR shortness of breath and fever or chills |

\*U.S. Centers for Disease Control and Prevention (CDC) COVID-19-like illness (CLI) definition was used to guide early diagnostic testing strategies from 17 January 2020 – 08 March 2020
(https://emergency.cdc.gov/han/han00426.asp).<sup>†</sup>U.S. Centers for Disease Control and Prevention (CDC) list of symptoms that may indicate COVID-19 infection (https://www.cdc.gov/coronavirus/2019-ncov/symptoms-testing/symptoms.html). This symptom list was last updated on 13 May 2020.
<sup>‡</sup>U.S. Council of State and Territorial Epidemiologists (CSTE) original clinical criteria for COVID-19 reporting (https://cdn.ymaws.com/www.cste.org/resource/resmgr/2020ps/interim-20-id-01_covid-19.pdf). This interim position statement (Interim-20-ID-01) was approved on 05 April 2020 and was replaced by Interim-20-ID-02 on 07 August 2020.
<sup>§</sup>U.S. Council of State and Territorial Epidemiologists (CSTE) revised clinical criteria for COVID-19 reporting (https://cdn.ymaws.com/www.cste.org/resource/resmgr/ps/positionstatement2020/interim-20-id-02_COVID-19.pdf). This interim position statement (Interim-20-ID-02) was approved on 07 August 2020 and replaced Interim-20-ID-01.
<sup>□</sup>Influenza-like illness (ILI) outpatient visit information collected through the U.S. Outpatient Influenza-like Illness Surveillance Network (ILINet) (https://www.cdc.gov/flu/weekly/overview.htm#anchor_1539281266932). This collaborative effort between CDC, state and local health departments, and healthcare providers has been tracking patients with ILI since the 1997-1998 influenza season.
<sup>¶</sup>World Health Organization (WHO) acute respiratory infection (ARI) definition for community-based respiratory syncytial virus (RSV) surveillance (https://www.who.int/influenza/rsv/rsv_case_definition/en/). Last updated 04 February 2020.
<sup>\*\*</sup>Compound symptom combinations were derived from all symptoms recorded at any time prior to enrollment (and after the index case's symptom onset date) through the end of the 14-day observation period.

We aimed to describe the diagnostic performance of two existing case definitions for COVID-19, the CDC COVID-19 symptom list, and two longstanding viral respiratory disease surveillance definitions among persons with confirmed SARS-CoV-2 exposure, stratified between adults and children. We also aimed to derive novel, practical symptom combinations in the same population. We interpreted the results primarily within the framework of two core public health surveillance functions: 1) symptom-based screening of individuals to guide SARS-CoV-2 diagnostic testing, contact tracing, and community-based isolation and quarantine, and 2) estimating disease frequency in persons with documented SARS-CoV-2 exposure. For symptom screening, we considered the merits of novel combinations when unlimited, timely diagnostic testing is unavailable.

## Methods

### Study Design and Data Collection

CDC collaborated with state and local health departments in the Milwaukee, Wisconsin and Salt Lake City, Utah metropolitan areas in the United States to identify and enroll a convenience sample of people with laboratory-confirmed SARS-CoV-2 infection and their household contacts from 22 March to 22 April 2020. Ours are secondary analyses of this household transmission investigation whose methods were previously published in detail.^15^ This activity was reviewed by CDC and was conducted consistent with applicable federal law and CDC policy. See e.g., 45 C.F.R. part 46, 21 C.F.R. part 56; 42 U.S.C. §241(d);□5 U.S.C. §552a; 44 U.S.C. §3501 et seq.

We administered questionnaires to household contacts to assess the presence of 15 symptoms during the 14 days prior to or at enrollment (day 0). Additionally, participants completed a daily symptom diary during days 1⍰14 after enrollment. We collected serum and upper respiratory specimens (i.e., both nasopharyngeal [NP] and anterior nares swabs) on day 0 and day 14. We additionally collected NP swabs at any interim date if any household contact newly developed or had worsening of any one of 15 symptoms consistent with COVID-19: nasal congestion or runny nose, sore throat, cough, chest pain, shortness of breath, discomfort while breathing, wheezing, headache, new loss of taste or smell, fever/chills, fatigue, muscle aches, diarrhea (≥3 loose stools per day), abdominal pain, or nausea/vomiting. The Milwaukee Health Department and Utah Public Health Laboratories tested the swabs using the CDC real-time Reverse Transcriptase Polymerase Chain Reaction (RT-PCR) assay for SARS-CoV-2,^16^ and CDC tested sera using a CDC-developed SARS-CoV-2 enzyme-linked immunosorbent assay (ELISA).^17^

### Definitions

We defined a household contact to be a COVID-19 case if they had at least one specimen test positive for SARS-CoV-2 by RT-PCR. We classified persons <18 years of age as children, and persons ≥18 years of age as adults. We combined all symptoms recorded at any time prior to enrollment (and after the index case’s symptom onset date) through the end of the 14-day observation period. We assessed individual symptoms, existing symptom combinations, and newly constructed symptom combinations for their association with SARS-CoV-2 test result by RT-PCR (Table 1). We asked enrollees to state whether they experienced any loss of taste and, separately, smell during the specified time period. For enrollees who responded yes to this question, we then asked whether the loss was partial or complete. We defined loss and/or dysfunction of taste or smell to include any level of loss, whether partial or complete. For ARI, we interpreted coryza as runny nose or nasal congestion.

### Analytic Methods

We excluded household contacts from the main analysis if not present at enrollment or not completing the study procedures. Our analysis of combinations predictive of COVID-19 included all 15 symptoms surveyed. We formally described the diagnostic performance of each individual symptom, existing COVID-19 case definitions, respiratory illness case definitions, and newly constructed symptom combinations (Table 1). The goal of assessing symptom combinations was to accurately divide the population into two groups: those who tested positive for SARS-CoV-2 and those who tested negative. For a given combination, we calculated the association of the combination with respect to laboratory-confirmed SARS-CoV-2 infection, yielding the number of contacts who were true positive (TP) (i.e., positive symptom profile and positive test), false negative (FN) (i.e., negative symptom profile and positive test), false positive (FP) (i.e., positive symptom profile and negative test), or true negative (TN) (i.e., negative symptom profile and negative test). From these values, we calculated the symptom combination’s sensitivity (i.e., TP / [TP + FN]), specificity (i.e., TN / [TN + FP]), positive predictive value (PPV) (i.e., TP / [TP + FP]), negative predictive value (NPV) (i.e., TN / [TN + FN]), F⍰1 score (the harmonic mean of sensitivity and PPV), and Youden’s index ([sensitivity + specificity] ⍰ 100). To determine how well each definition would estimate prevalence in a syndromic surveillance system, we also calculated the difference in the number of positive symptom screens (i.e., TP + FP) from the actual number of contacts who tested positive by RT-PCR. We assessed combinations across all ages and in children and adults separately, and we reported all measures on the percentage scale. To assess variability in each performance measure, we constructed bias-corrected and accelerated bootstrap confidence intervals^18^ over 10,000 pseudosamples constructed by resampling households with replacement. We reported 95% confidence intervals with two exceptions. For measures estimated at 100% in the observed sample, we omitted confidence intervals, because the pseudosamples could not exhibit any variability. For the difference in specificity and sensitivity between adults and children, we reported 97·5% confidence intervals (a Bonferroni correction) to allow for a 95% joint confidence level regarding the differences in each pair.

We adapted innovative methods previously applied in the low-resource context to derive a parsimonious symptom combination to prioritize diagnostic testing for tuberculosis.^19^ We chose this approach to be as comprehensive as practical for COVID-19 in that it systematically assessed nearly every conceivable combination of symptoms.

First, we searched over 245,000 combinations of between one and 15 symptoms (i.e., simple combinations of the form “at least *m* symptoms present out of *n* symptoms considered”). We gave greater weight to combinations with high F—1 score or high Youden’s index. We then conducted an exhaustive search using pairs of these “m—of—n” combinations (i.e., compound combinations) to allow for more nuanced combinations. We limited this second search to single combinations of no more than five symptoms, such that the number of total symptoms evaluated for a compound combination was never more than ten. We allowed each pair of combinations to be joined by the logical operators [AND] and [OR], yielding approximately 73 million unique combination pairs. After the search, we selected four combination pairs to include in the primary analysis on the basis of diagnostic performance and parsimony. We measured diagnostic performance by F–1 score (higher being better). We measured parsimony by the total number of symptoms evaluated (fewer being better) (Table 1).

We performed all calculations in R 4·0·0 (R Core Team), Python 3·7 (Python Software Foundation), or both. To allow for parallel processing, the exhaustive combinatorial search and bootstrap confidence intervals were implemented on a scientific workstation with 24 logical cores and 64 GB of RAM. De-identified data and analytic scripts in R and Python are publicly available through a GitHub repository: https://github.com/scotthlee/covid-casedefs.

## Findings

### Study Population

We enrolled 199 contacts of index patients with laboratory-confirmed SARS-CoV-2 infections within 62 households. We excluded one contact who was not living in the home on the day of enrollment, one who was hospitalized at enrollment, and two who did not consent to have specimens collected. Ten contacts had negative RT-PCR and positive serology test results; they were also excluded from the primary analyses. Therefore, our analyses included the remaining 185 household contacts. The median time interval from index patient’s symptom onset date to enrollment was 10 days (interquartile range [IQR]: 7⍰13). About half (95; 51%) were female. 108 (58%) were Caucasian/white, 32 (17%) Latinx/Hispanic, 23 (12%) African American/black, 14 (8%) Asian, four (2%) Native American, and four (2%) multiracial. The median age was 22 years ([IQR]: 14⍰47), with 122 (66%) adults and 63 (34%) children. Among children, nine (14%) were <5 years, 19 (30%) were 5⍰9 years, and 35 (56%) were 10⍰17 years of age. SARS-CoV-2 infection was detected by RT-PCR in 49 (27%) household contacts. Separated by age group, 35/122 (29%) adults and 14/63 (22%) children had laboratory-confirmed SARS-CoV-2 infection by RT-PCR. Among the 49 RT-PCR-positive contacts, most (45; 92%) also had a positive serology result, three had a negative serology result, and one was not tested by serology.

### Performance Characteristics for Individual Symptoms (All Ages Pooled)

Individual symptoms with the highest sensitivity were nasal congestion or rhinorrhea, headache, and cough (Table 2, Figure 1). Many of the individual symptoms reported were highly specific, although generally resulting in lower sensitivity. The exception was loss or dysfunction of taste or smell (categorized as a single symptom), which had a moderate sensitivity of 63% (95% CI 47⍰77%), high specificity (96%; 95% CI 90⍰99%), high NPV (88%; 95% CI 80⍰93%), and the highest PPV (84%; 95% CI 64⍰94%), Youden’s index (59%; 95% CI 42⍰73%), and F⍰1 score (72%; 95% CI 57⍰83%).

**Table 2.** Diagnostic performance indicators for individual COVID-19 symptoms, existing case definitions, and derived compound symptom combinations for a community cohort of 185 people with household COVID-19 exposure in Utah and Wisconsin, United States, March–May 2020

|  | SARS-CoV-2-negative<br>(n=136) |  | SARS-CoV-2-positive<br>(n=49) |  | Specificity* |  | Sensitivity<sup>†</sup> |  | NPV<sup>‡</sup> |  | PPV<sup>§</sup> |  | Youden's index<sup>□</sup> |  | F-1 score<sup>¶</sup> |  | Difference in prevalence** |  |
| --- | --- | --- | --- | --- | --- | --- | --- | --- | --- | --- | --- | --- | --- | --- | --- | --- | --- | --- |
|  | TN<sup>††</sup> | FP<sup>‡‡</sup> | FN<sup>§§</sup> | TP<sup>□□</sup> | Value | (95% CI) | Value | (95% CI) | Value | (95% CI) | Value | (95% CI) | Value | (95% CI) | Value | (95% CI) | Value | (95% CI) |
| **Individual symptoms** |  |  |  |  |  |  |  |  |  |  |  |  |  |  |  |  |  |  |
| *Upper respiratory* |  |  |  |  |  |  |  |  |  |  |  |  |  |  |  |  |  |  |
| Nasal congestion or rhinorrhea | 66 | 70 | 5 | 44 | 49 | (38, 59) | 90 | (80, 96) | 93 | (85, 97) | 39 | (27, 50) | 38 | (26, 50) | 54 | (42, 65) | 133 | (78, 227) |
| Sore throat | 96 | 40 | 22 | 27 | 71 | (61, 79) | 55 | (40, 68) | 81 | (72, 88) | 40 | (26, 56) | 26 | (10, 40) | 47 | (33, 59) | 37 | (0, 98) |
| *Lower respiratory* |  |  |  |  |  |  |  |  |  |  |  |  |  |  |  |  |  |  |
| Cough | 103 | 33 | 13 | 36 | 76 | (65, 84) | 74 | (60, 85) | 89 | (81, 94) | 52 | (37, 67) | 49 | (36, 62) | 61 | (49, 72) | 41 | (8, 94) |
| Chest pain | 122 | 14 | 35 | 14 | 90 | (83, 94) | 29 | (15, 42) | 78 | (69, 85) | 50 | (24, 70) | 18 | (5, 33) | 36 | (21, 51) | -43 | (-62, -15) |
| Shortness of breath | 125 | 11 | 36 | 13 | 92 | (86, 96) | 27 | (15, 40) | 78 | (67, 85) | 54 | (31, 75) | 18 | (6, 32) | 36 | (22, 49) | -51 | (-68, -24) |
| Discomfort while breathing | 132 | 4 | 37 | 12 | 97 | (93, 99) | 25 | (14, 39) | 78 | (69, 85) | 75 | (40, 92) | 22 | (10, 36) | 37 | (22, 54) | -67 | (-80, -49) |
| Wheezing | 133 | 3 | 44 | 5 | 98 | (94, 99) | 10 | (4, 21) | 75 | (65, 83) | 63 | (20, 100) | 8 | (1, 19) | 18 | (7, 33) | -84 | (-92, -68) |
| *Neurological* |  |  |  |  |  |  |  |  |  |  |  |  |  |  |  |  |  |  |
| Headache | 86 | 50 | 7 | 42 | 63 | (52, 73) | 86 | (75, 93) | 93 | (85, 97) | 46 | (33, 59) | 49 | (35, 60) | 60 | (47, 70) | 88 | (44, 160) |
| Taste and/or smell dysfunction | 130 | 6 | 18 | 31 | 96 | (90, 99) | 63 | (47, 77) | 88 | (80, 93) | 84 | (64, 94) | 59 | (42, 73) | 72 | (57, 83) | -25 | (-43, -5) |
| *Constitutional* |  |  |  |  |  |  |  |  |  |  |  |  |  |  |  |  |  |  |
| Fever or chills | 113 | 23 | 18 | 31 | 83 | (75, 89) | 63 | (49, 76) | 86 | (78, 92) | 57 | (40, 72) | 46 | (30, 60) | 60 | (46, 72) | 10 | (-15, 51) |
| Fatigue | 94 | 42 | 20 | 29 | 69 | (59, 79) | 59 | (41, 74) | 83 | (74, 89) | 41 | (25, 57) | 28 | (10, 44) | 48 | (33, 63) | 45 | (8, 106) |
| Myalgia | 116 | 20 | 21 | 28 | 85 | (77, 91) | 57 | (42, 70) | 85 | (77, 90) | 58 | (41, 74) | 42 | (28, 56) | 58 | (44, 69) | -2 | (-28, 37) |
| *Gastrointestinal* |  |  |  |  |  |  |  |  |  |  |  |  |  |  |  |  |  |  |
| Diarrhea | 108 | 28 | 31 | 18 | 79 | (69, 88) | 37 | (20, 55) | 78 | (66, 86) | 39 | (22, 59) | 16 | (-3, 36) | 38 | (23, 54) | -6 | (-40, 48) |
| Abdominal pain | 120 | 16 | 34 | 15 | 88 | (80, 94) | 31 | (19, 45) | 78 | (68, 85) | 48 | (27, 73) | 19 | (4, 35) | 38 | (23, 53) | -37 | (-59, -2) |
| Nausea | 124 | 12 | 40 | 9 | 91 | (85, 95) | 18 | (10, 31) | 76 | (65, 83) | 43 | (20, 67) | 10 | (-1, 22) | 26 | (14, 40) | -57 | (-74, -30) |
| **Existing case definitions** |  |  |  |  |  |  |  |  |  |  |  |  |  |  |  |  |  |  |
| *COVID-19 and respiratory illness surveillance case definitions* |  |  |  |  |  |  |  |  |  |  |  |  |  |  |  |  |  |  |
| CDC symptom list<sup>¶¶</sup> | 40 | 96 | 0 | 49 | 29 | (17, 45) | 100 | NA<sup>□□□□</sup> | 100 | NA | 34 | (24, 45) | 29 | (17, 45) | 51 | (38, 62) | 196 | (122, 322) |
| ARI<sup>***</sup> | 52 | 84 | 2 | 47 | 38 | (26, 51) | 96 | (86, 100) | 96 | (88, 100) | 36 | (25, 48) | 34 | (23, 45) | 52 | (40, 64) | 167 | (102, 279) |
| CSTE combination 1<sup>†††</sup> | 79 | 57 | 3 | 46 | 58 | (47, 69) | 94 | (82, 98) | 96 | (91, 99) | 45 | (32, 58) | 52 | (39, 63) | 61 | (47, 72) | 110 | (64, 190) |
| CSTE combination 2<sup>‡‡‡</sup> | 59 | 77 | 1 | 48 | 43 | (35, 52) | 98 | (88, 100) | 98 | (90, 100) | 38 | (30, 47) | 41 | (32, 50) | 55 | (46, 64) | 155 | (110, 225) |
| CLI<sup>§§§</sup> | 93 | 43 | 7 | 42 | 68 | (57, 79) | 86 | (70, 94) | 93 | (88, 97) | 49 | (35, 64) | 54 | (40, 65) | 63 | (48, 75) | 74 | (35, 141) |
| ILI<sup>□□□</sup> | 122 | 14 | 24 | 25 | 90 | (83, 94) | 51 | (35, 65) | 84 | (75, 90) | 64 | (45, 79) | 41 | (25, 55) | 57 | (42, 69) | -20 | (-41, 9) |
| **Derived compound symptom combinations<sup>¶¶¶</sup>** |  |  |  |  |  |  |  |  |  |  |  |  |  |  |  |  |  |  |
| Combination 1<sup>****</sup> | 129 | 7 | 12 | 37 | 95 | (89, 98) | 76 | (60, 87) | 92 | (85, 96) | 84 | (67, 94) | 70 | (54, 82) | 80 | (67, 88) | -10 | (-28, 9) |
| Combination 2<sup>††††</sup> | 125 | 11 | 10 | 39 | 92 | (84, 97) | 80 | (65, 89) | 93 | (86, 96) | 78 | (59, 91) | 72 | (56, 83) | 79 | (66, 88) | 2 | (-17, 30) |
| Combination 3<sup>‡‡‡‡</sup> | 126 | 10 | 11 | 38 | 93 | (85, 97) | 78 | (63, 88) | 92 | (86, 96) | 79 | (60, 91) | 70 | (54, 82) | 78 | (66, 87) | -2 | (-20, 24) |
| Combination 4<sup>§§§§</sup> | 129 | 7 | 13 | 36 | 95 | (89, 98) | 74 | (59, 84) | 91 | (84, 95) | 84 | (67, 94) | 68 | (54, 80) | 78 | (67, 86) | -12 | (-29, 8) |

\*Specificity is the probability of testing negative when disease is absent.
<sup>†</sup>Sensitivity is the probability of testing positive when disease is present.
<sup>‡</sup>NPV = negative predictive value. NPV is the probability of a patient not having disease when test is negative.
<sup>§</sup>PPV = positive predictive value. PPV is the probability of a patient having disease when test is positive.
<sup>□</sup>Youden's index is defined as sensitivity plus specificity minus 100 (perfect score=100).
<sup>¶</sup>F-1 score is defined as the harmonic mean of sensitivity and PPV (perfect score=100).
<sup>\*\*</sup>Difference in prevalence is defined as the difference in the number of positive symptom screens (i.e., true positive plus false positive) for each combination from the actual number of contacts who tested positive by RT-PCR.
<sup>††</sup>TN = true negative.
<sup>‡‡</sup>FP = false positive.
<sup>§§</sup>FN = false negative.
<sup>□□</sup>TP = true positive.
<sup>¶¶</sup>U.S. Centers for Disease Control and Prevention (CDC) list of symptoms that may indicate COVID-19 infection (https://www.cdc.gov/coronavirus/2019-ncov/symptoms-testing/symptoms.html). This symptom list was last updated on 13 May 2020.
<sup>†††</sup>U.S. Council of State and Territorial Epidemiologists (CSTE) revised clinical criteria for COVID-19 reporting (https://cdn.ymaws.com/www.cste.org/resource/resmgr/ps/positionstatement2020/interim-20-id-02_COVID-19.pdf). This interim position statement (Interim-20-ID-02) was approved on 07 August 2020 and replaced Interim-20-ID-01.
<sup>§§§</sup>U.S. Centers for Disease Control and Prevention (CDC) COVID-19-like illness (CLI) definition was used to guide early diagnostic testing strategies from 17 January 2020 – 08 March 2020 (https://emergency.cdc.gov/han/han00426.asp).

**Figure 1.**
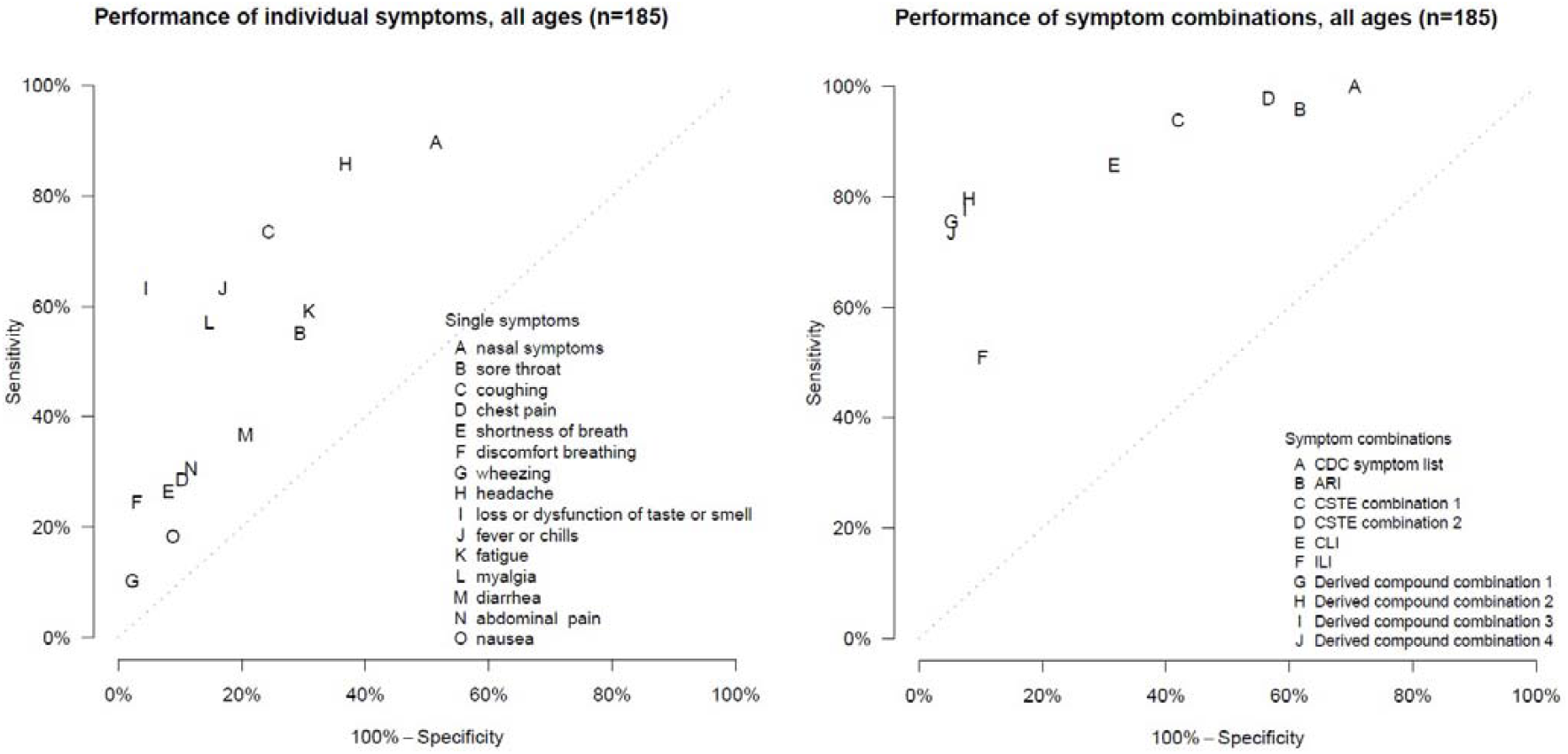
Sensitivity and 100%-specificity for individual COVID-19 symptoms, existing case definitions, and derived compound symptom combinations for a community cohort of 185 individuals with household exposure to COVID-19 in Utah and Wisconsin, United States, March–May 2020. *Specificity is the probability of testing negative when disease is absent. ^†^Sensitivity is the probability of testing positive when disease is present. ^‡^U.S. Centers for Disease Control and Prevention (CDC) list of symptoms that may indicate COVID-19 infection (https://www.cdc.gov/coronavirus/2019-ncov/symptoms-testing/syptoms.html). This symptom list was last updated on 13 May 2020. ^§^World Health Organization (WHO) acute respiratory infection (ARI) definition for community-based respiratory syncytial virus (RSV) surveillance (https://www.who.int/influenza/rsv/rsv_case_definition/en/). Last updated 04 February 2020. ^□^U.S. Council of State and Territorial Epidemiologists (CSTE) original clinical criteria for COVID-19 reporting (https://cdn.ymaws.com/www.cste.org/resource/resmgr/2020ps/interim-20-id-01_covid-19.pdf). This interim position statement (Interim-20-ID-01) was approved on 05 April 2020 and was replaced by Interim-20-ID-02 on 07 August 2020. ^¶^U.S. Council of State and Territorial Epidemiologists (CSTE) revised clinical criteria for COVID-19 reporting (https://cdn.ymaws.com/www.cste.org/resource/resmgr/ps/positionstatement2020/interim-20-id-02_COVID-19.pdf). This interim position statement (Interim-20-ID-02) was approved on 07 August 2020 and replaced Interim-20-ID-01. **U.S. Centers for Disease Control and Prevention (CDC) COVID-19-like illness (CLI) definition was used to guide early diagnostic testing strategies from 17 January 2020 – 08 March 2020 (https://emergency.cdc.gov/han/han00426.asp). ^††^Influenza-like illness (ILI) outpatient visit information collected through the U.S. Outpatient Influenza-like Illness Surveillance Network (ILINet) (https://www.cdc.gov/flu/weekly/overview.htm#anchor_1539281266932). This collaborative effort between CDC, state and local health departments, and healthcare providers has been tracking patients with ILI since the 1997-1998 influenza season. ^‡‡^Combination 1: Taste and/or smell dysfunction, OR one of the following: shortness of breath, myalgia, or fever or chills. ^§§^Combination 2: Taste and/or smell dysfunction or discomfort breathing, OR at least two of the following: shortness of breath, wheezing, or fever/chills. ^□□^Combination 3: Taste and/or smell dysfunction, OR at least two of the following: shortness of breath, wheezing, discomfort breathing, or fever/chills. ^¶¶^Combination 4: Taste and/or smell dysfunction, OR shortness of breath and fever/chills. Points closest to the upper left corner represent those with the highest sensitivity and specificity values.

### Performance Characteristics for Existing COVID-19 Case Definitions, the CDC Symptom List, and Respiratory Syndromic Surveillance Networks (All Ages Pooled)

Among the existing case definitions, sensitivity was perfect (100%) for the CDC symptom list definition, and also high for ARI (96%; 95% CI 86⍰100%), CSTE combination 1 (original); 94%; 95% CI 82⍰98%), CSTE combination 2 (revised); 98%; 95% CI 88⍰100%), and CLI (86%; 95% CI 70⍰94%) (Table 2, Figure 1). While these definitions offered high sensitivity, they were poorly specific. Conversely, ILI was highly specific (90%; 95% CI 83⍰94%) but insensitive (51%; 95% CI 35⍰65%). All existing definitions demonstrated low PPV. Youden’s indices and F⍰1 scores were highest for CSTE combination 1 and CLI, though still suboptimal. None of the existing definitions predicted prevalence well; the difference from true prevalence ranged from ⍰20 for ILI to 196 for the CDC symptom list. Compared to CSTE combination 1, CSTE combination 2 had slightly higher sensitivity and NPV, but performed more poorly on all other diagnostic performance indicators.

### Performance Characteristics for Derived Compound Symptom Combinations (All Ages Pooled)

The four highest performing novel symptom combinations, based on F⍰1 score and parsimony, were compound symptom combinations that included dysfunction of taste or smell. These four combinations performed similarly well on all performance measures (Table 2, Figure 1). We determined compound symptom combination 3 (i.e., loss or dysfunction of taste or smell, or at least two of the following: shortness of breath, wheezing, discomfort breathing, or fever/chills), to be simple to implement, have higher specificity (93%; 95% CI 85⍰97%), NPV (92%; 95% CI 86⍰96%), PPV (79%; 95% CI 60⍰91%), Youden’s index (70%; 95% CI 54⍰82%), and F⍰1 score (78%; 95% CI 66⍰87%) than existing case definitions. The compound symptom combination 3 showed near-perfect prevalence prediction (⍰2; 95% CI ⍰20⍰24), and sensitivity was moderately high (78%; 95% CI 63⍰88%).

### Adult-Child Differences in Discriminatory Performance

The accuracy of symptom profiles for defining RT-PCR confirmed COVID-19 differed by age (Table 3, Figure 2). Overall, existing case definitions were less sensitive in children compared to adults. One exception, the CDC symptom list for priority testing (Table 1), captured all COVID-19 cases regardless of age. The existing case definitions were more specific in children, but the greater specificity was statistically significant for CSTE combination 1 only. Individual symptoms showed a similar pattern of lower sensitivity among children, notably taste/smell dysfunction. Sore throat was more sensitive in children, and fever/chills and nausea were similar regardless of age group. We observed a similar pattern of increased specificity for most derived symptom combinations in children (Table 3, Figure 2). Cough was the sole symptom where the difference in both sensitivity and specificity was statistically significant. For both children and adults, the CLI case definition provided the greatest balance between both sensitivity and specificity (Youden’s Index 53%; 95% CI 8⍰80% vs. 52%; 95% CI 36⍰66%, respectively) and harmonization of sensitivity with PPV (F⍰1 61%; 95% CI 26⍰83% vs. 63%; 95% CI 49⍰76%, respectively) (Table 2). CLI also most accurately predicted overall prevalence amongst children (percent difference from true prevalence 36%; 95% CI ⍰17⍰157) (Table 3).

**Table 3.**
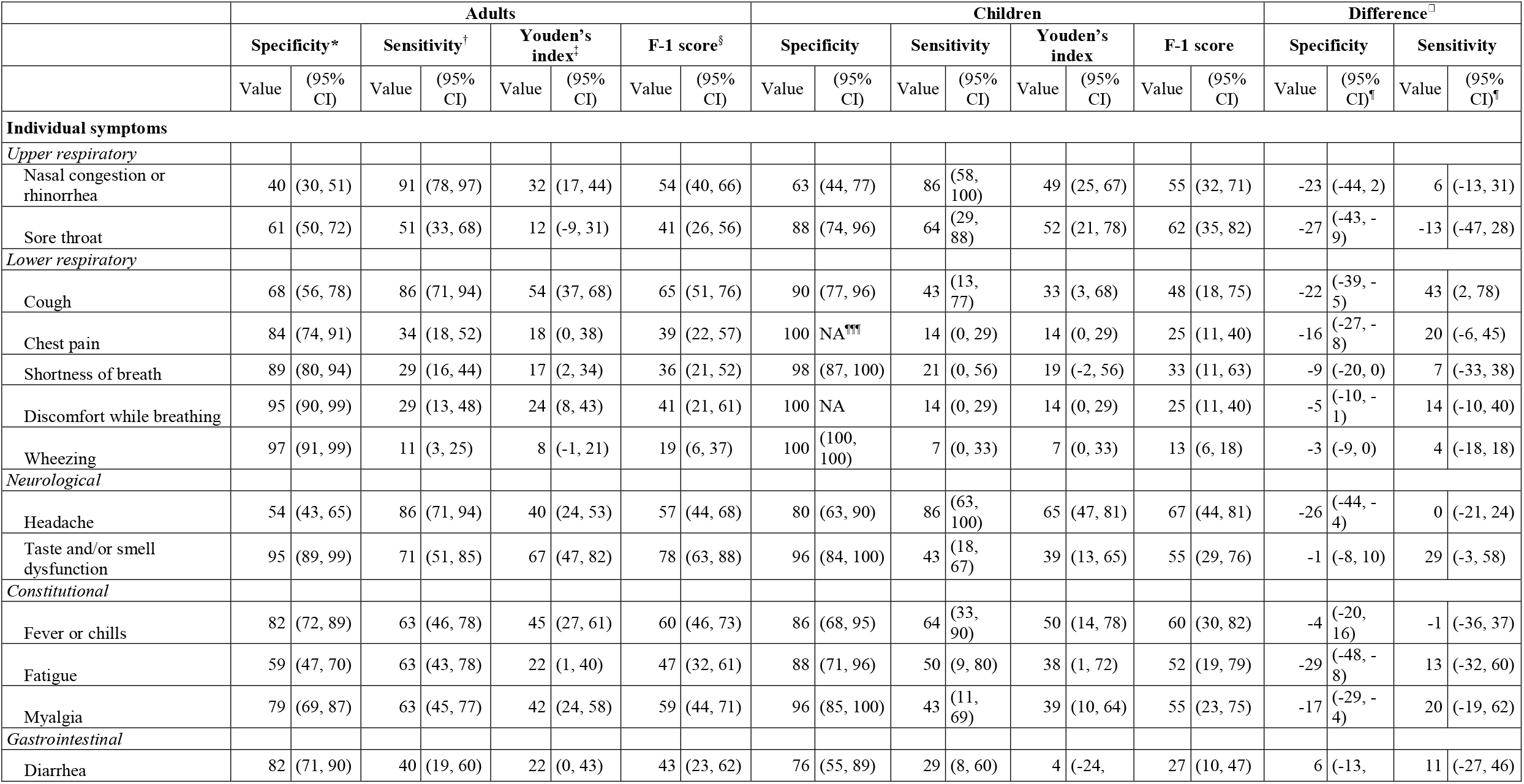
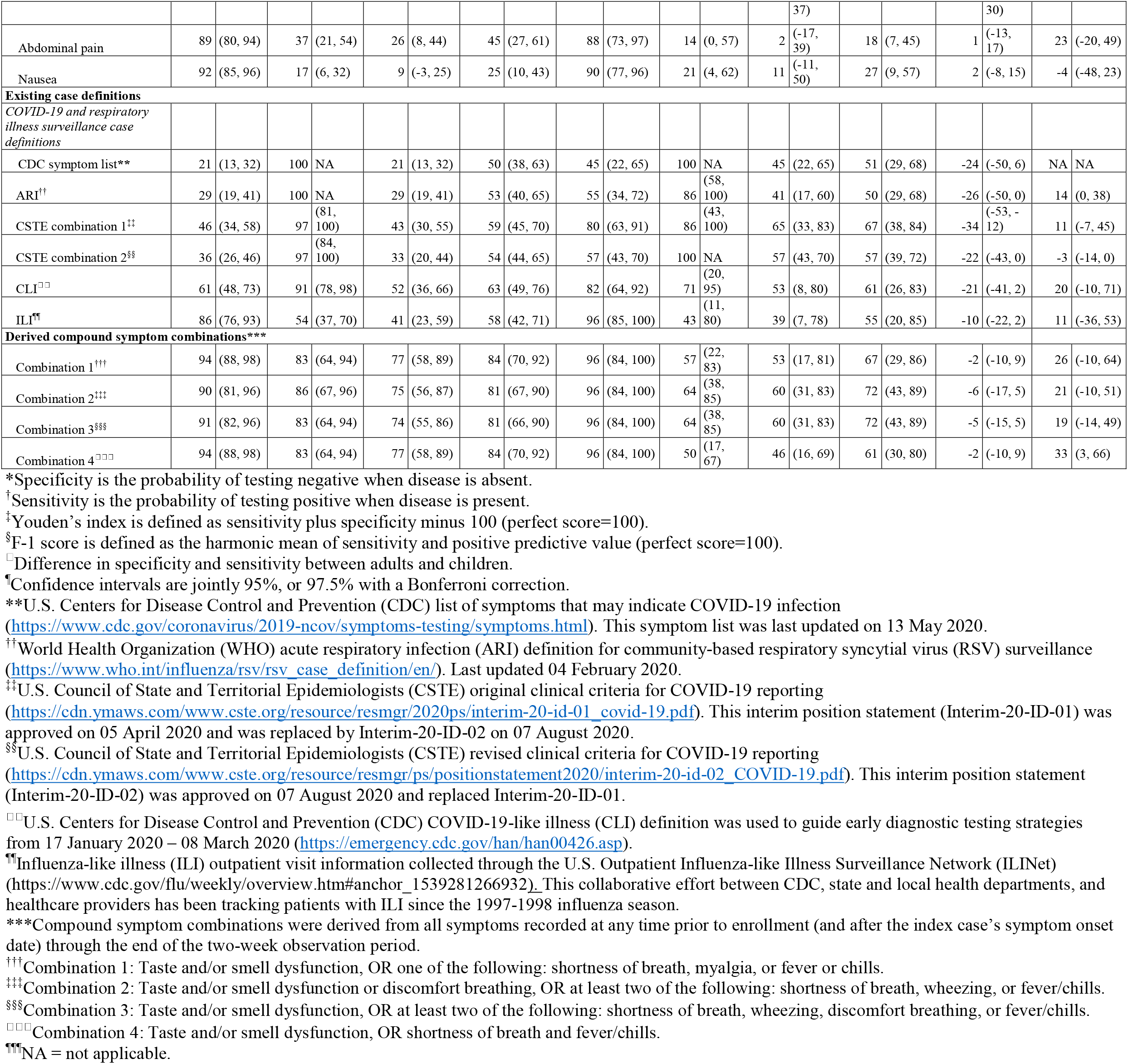
Diagnostic performance indicators for individual COVID-19 symptoms, existing case definitions, and derived compound symptom combinations by age group for 122 adults and 63 children with household exposure to COVID-19 in Utah and Wisconsin, United States, March– May 2020

**Figure 2.**
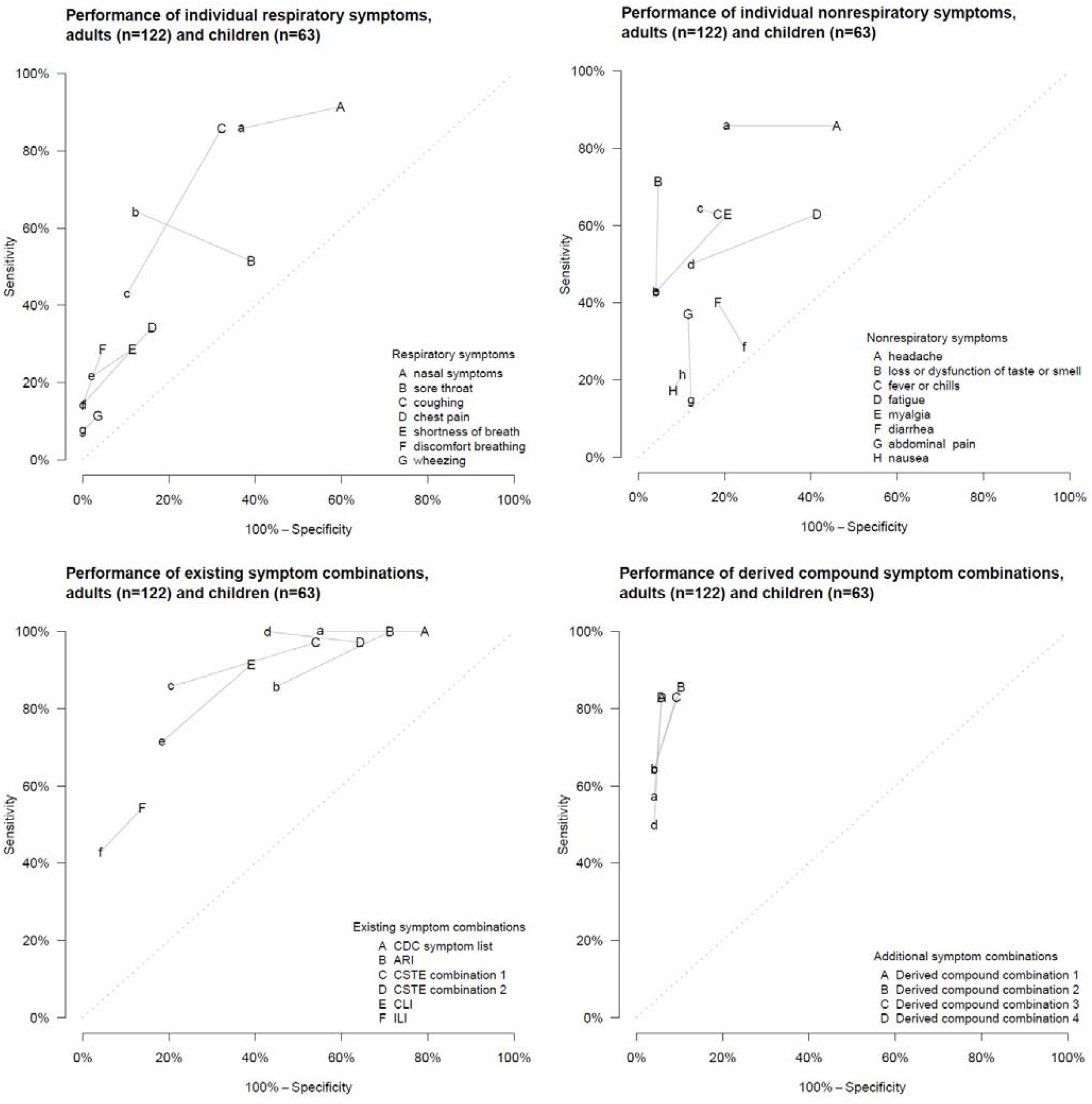
Sensitivity and 100%-specificity for individual COVID-19 symptoms, existing case definitions, and derived compound symptom combinations for a community cohort of 122 adults (upper case letters) and 63 children (lower case letters) with household exposure to COVID-19 in Utah and Wisconsin, United States, March–May 2020. ^*^Specificity is the probability of testing negative when disease is absent. ^†^Sensitivity is the probability of testing positive when disease is present. ^‡^U.S. Centers for Disease Control and Prevention (CDC) list of symptoms that may indicate COVID-19 infection (https://www.cdc.gov/coronavirus/2019-ncov/symptoms-testing/symptoms.html). This symptom list was last updated on 13 May 2020. ^§^World Health Organization (WHO) acute respiratory infection (ARI) definition for community-based respiratory syncytial virus (RSV) surveillance (https://www.who.int/influenza/rsv/rsv_case_definition/en/). Last updated 04 February 2020. ^□^U.S. Council of State and Territorial Epidemiologists (CSTE) clinical criteria for COVID-19 reporting (https://cdn.ymaws.com/www.cste.org/resource/resmgr/2020ps/interim-20-id-01_covid-19.pdf). This interim position statement (Interim-20-ID-01) was approved on 05 April 2020 and was replaced by Interim-20-ID-02 on 07 August 2020. ^¶^U.S. Council of State and Territorial Epidemiologists (CSTE) clinical criteria for COVID-19 reporting (https://cdn.ymaws.com/www.cste.org/resource/resmgr/ps/positionstatement2020/interim-20-id-02_COVID-19.pdf). This interim position statement (Interim-20-ID-02) was approved on 07 August 2020 and replaced Interim-20-ID-01. **U.S. Centers for Disease Control and Prevention (CDC) COVID-19-like illness (CLI) definition was used to guide early diagnostic testing strategies from 17 January 2020 – 08 March 2020 (https://emergency.cdc.gov/han/han00426.asp). ^††^Influenza-like illness (ILI) outpatient visit information collected through the U.S. Outpatient Influenza-like Illness Surveillance Network (ILINet) (https://www.cdc.gov/flu/weekly/overview.htm#anchor_1539281266932). This collaborative effort between CDC, state and local health departments, and healthcare providers has been tracking patients with ILI since the 1997-1998 influenza season. ^‡‡^Combination 1: Taste and/or smell dysfunction, OR one of the following: shortness of breath, myalgia, or fever or chills. ^§§^Combination 2: Taste and/or smell dysfunction or discomfort breathing, OR at least two of the following: shortness of breath, wheezing, or fever/chills. ^□□^Combination 3: Taste and/or smell dysfunction, OR at least two of the following: shortness of breath, wheezing, discomfort breathing, or fever/chills. ^¶¶^Combination 4: Taste and/or smell dysfunction, OR shortness of breath and fever/chills. Points closest to the upper left corner represent those with the highest sensitivity and specificity values.

#### Interpretation

Existing case surveillance definitions for COVID-19, as shown in Table 1, were generally sensitive in our study conducted among household contacts of infected persons, a population with proven SARS-CoV-2 exposure.

However, they tended to have low specificity and poorly estimated disease prevalence. By systematically screening novel definitions that optimized sensitivity, specificity, and PPV, we improved community prevalence estimation and overall accuracy of individual screening, which could be useful if diagnostic testing is limited. In particular, we affirmed loss or dysfunction of taste or smell as a uniquely discerning characteristic central to constructing an effective, concise case surveillance definition when applied across all age groups (i.e., derived compound combination 3).

An appropriate discriminatory balance between sensitivity and specificity for a newly emerging pathogen depends on the objectives of the surveillance activity.^20^ Highly sensitive case definitions capture a larger proportion of true COVID-19 cases, which is ideal when diagnostic testing is widely available and results are timely. Highly sensitive definitions, however, generally rule in a larger number of non-cases (i.e., FP symptom screens).^21^ In addition to testing resources, the public health system’s tolerance for false-positive screens is, of course, dependent on human resources. This is especially apparent when intensive interventions involve extensive contact tracing, isolation and quarantine. At high community COVID-19 prevalence, these intensive mitigation efforts may benefit from evidence-based prioritization. By example, CSTE combination 2 had a FP symptom screening rate (77/136; 57%) eight times that for derived compound combination 3 (10/136; 7%) in our cohort. At the population level, such differences could expose shortcomings in resources for core interventions, such as universal contact tracing. The COVID-19 response has repeatedly been strained in these requisite areas.^22-24^ Novel symptom screening criteria that more tightly couple sensitivity and specificity (i.e., diagnostic accuracy), such as the derived compound combination 3, could help to prioritize interventions when strategically deployed. This principle may also apply when evaluating novel vaccines or therapeutics in large clinical trials involving thousands of participants, where feasibility constraints often dictate the use of symptom-prioritized testing to confirm outcomes. Still, highly sensitive symptom rules, such as CSTE combination 2, are preferred for COVID-19 when resources are unlimited.

For using syndromic surveillance systems to estimate community burden, the highly sensitive existing case definitions overestimated true burden. Conversely, highly specific case definitions, such as ILI, may detect changes in disease trends over time but underestimate true burden.^20^ ILI underestimated disease prevalence by more than 80% in this study population. Current laboratory-based surveillance grossly under-ascertains incidence,^25^ especially where diagnostic testing is not easily accessible or widespread. Retailoring community-based syndromic surveillance systems already in place^26^ (i.e., altering the symptoms included or applying a correction factor based on results such as ours) would more accurately reflect true burden.

For most symptoms and their combinations, overall performance, most notably sensitivity, differed between children and adults. These findings are consistent with prior observations whereby children generally show fewer and milder symptoms of COVID-19 compared with adults,^27^ and COVID-19 syndromes vary across ages.^13^ The small number of children with COVID-19 in this cohort limits the conclusion of specific recommendations, but further examination into the utility of age-specific case definitions is warranted in considering policies for schools and other child congregate settings, and for deriving accurate burden estimates from syndromic surveillance.

While the number of individuals in this study is relatively small, our investigation is among the largest and most well-characterized of its kind to date. We collected extensive symptom data, which yielded a comprehensive assessment of multiple symptom combinations. We also employed inclusion criteria that were not based on disease status or symptom status, and a reference category based on standardized laboratory testing. Nonetheless, we acknowledge this study’s limitations. These analyses were not intended to produce definitive symptom combinations to be applied to the general public, however they may be used to guide the development of future candidate case definitions. One key consideration for future validation efforts is that enrollment started immediately after the precipitous decline in laboratory-confirmed influenza virus infections in the United States in mid-March 2020.^28^ Thus, our estimates of diagnostic performance may differ during the viral respiratory season. In addition, COVID-19 prevalence was higher for our study population (i.e., contacts of laboratory-confirmed household members) compared to the entire community, thereby limiting the generalizability of predictive values (although sensitivity and specificity remain unaffected by disease prevalence). Screening criteria applied to persons seeking medical care may also perform differently, as those individuals probably tend to have more severe illness.

Additionally, we showed that existing COVID-19 case definitions are highly sensitive and do well to screen in persons for testing and individual-level public health interventions like community isolation. In the first such endeavor for evaluating and deriving novel COVID-19 case surveillance definitions in a community setting among SARS-CoV-2—exposed individuals with largely mild illness, we evaluated novel symptom combinations for COVID-19 using methodology previously applied to tuberculosis in low resource settings.^19^ These derived combinations and CSTE definition 2 better estimated community disease burden and used taste and/or smell dysfunction as a primary component. The latter is supported by prior studies.^5,8-10^ Because most SARS-CoV-2 infections are mild^29^ and core public health functions may need prioritization when testing and other resources are limited, case definitions that accurately determine COVID-19 status in the general public may assist continued interruption of community transmission.^30^ When timely diagnostic testing is readily available, however, using less sensitive screening tools could inappropriately miss cases and lead to further community transmission.

Our study population, which includes persons across the age spectrum and enrolled participants independent of disease and symptom status, may better reflect the diagnostic performance in the general population than previously published research. Accurate clinical case definitions are likely to also apply to large clinical trials for candidate vaccines and therapeutics where serial confirmatory SARS-CoV-2 testing for any new symptom is impractical. It is important that our results be validated against the growing body of larger ambulatory surveillance databases in diverse communities and in other countries; in particular, our methodology should be assessed in the context of the annual influenza season, at varying community COVID-19 prevalence, and across the age spectrum. Such studies ideally can be accompanied by cost-effectiveness modeling of intervention strategies.

## Data Availability

De-identified data and analytic scripts in R and Python are publicly available through a GitHub repository: https://github.com/scotthlee/covid-casedefs.

**U.S. COVID-19 Household Investigation Team**

Michelle Banks, MS^*^, Katherine A. Battey, MPH^*^, Allison Binder, MPH^*^, Sean Buono, PhD^*,‡^, Rebecca J. Chancey, MD^*^, Ann Christiansen, MPH^**^, Erin E. Conners, PhD^*^, Trivikram Dasu, PhD^¶^, Patrick Dawson, PhD^*,†^, Elizabeth Dietrich, PhD^*^, Lindsey M. Duca, PhD^*,†^, Angela C. Dunn, MD^§^, Victoria L. Fields, DVM^*,†^, Garrett Fox, MPH^*^, Brandi D. Freeman, PhD^*,‡^, Radhika Gharpure, DVM^*,†^, Christopher Gregory, MD^*^, Tair Kiphibane, BSN^††^, Rebecca L. Laws, PhD^*^, Sandra Lester, PhD^*^, Nathaniel M. Lewis, PhD^*,†,§^, Perrine Marcenac, PhD^*,†^, Almea M. Matanock, MD^*^, Lisa Mills, PhD^*^, Henry Njuguna, MD^*^, Michelle O’Hegarty, PhD^*^, Daniel Owusu, DrPH^*,†^, Lindsey Page, MPH^¶^, Lucia Pawloski, PhD^*^, Eric Pevzner, PhD^*^, Mary Pomeroy, MSN^*,†^, Ian W. Pray, PhD^*,†,□^, Elizabeth M. Rabold, MD^*,†^, Jared R. Rispens, MD^*,†^, Phillip Salvatore, PhD^*,†^, Amy Schumacher, PhD^*^, Cuc H. Tran, PhD^*^, Jeni Vuong, MS^*^, Ashutosh Wadhwa, PhD^*,‡^, Ryan P. Westergaard, MD, PhD^**^, Sarah Willardson, MPH^‡‡^, Dongni Ye, PhD^*^, Sherry Yin, MPH^*^, Anna Yousaf, MD^*,†^

^*^COVID-19 Response Team, CDC

^†^Epidemic Intelligence Service, CDC

^‡^Laboratory Leadership Service, CDC

^§^Utah Department of Health

^□^Wisconsin Department of Health

^¶^City of Milwaukee Health Department

^**^North Shore (WI) Health Department

^††^Salt Lake City (UT) Health Department

^‡‡^Davis County (UT) Health Department

## Author Contributions

All authors contributed to study design, data collection, scientific interpretation, and review and approval of the final manuscript. In addition, the following authors made unique contributions:

Scott H. Lee and Charles M. Heilig conducted all data analysis, provided open analytic code for public dissemination, led data interpretation, wrote data-analytic methods, and assisted with integrating analysis and interpretation throughout the manuscript.

Robyn Atkinson-Dunn, Sanjib Bhattacharyya, Trivikram Dasu, Kim Christensen, Brandi D. Freeman, Sandra Lester, Lisa Mills, Lucia Pawloski, and Natalie J. Thornburg coordinated or performed diagnostic laboratory analyses.

Katherine A. Battey, Sean Buono, Rebecca J. Chancey, Ann Christiansen, Erin E. Connors, Patrick Dawson, Elizabeth Dietrich, Lindsey M. Duca, Victoria L. Fields, Radhika Gharpure, Christopher Gregory, Tair Kiphibane, Rebecca L. Laws, Nathaniel M. Lewis, Perrine Marcenac, Almea M. Matanock, Henry Njuguna, Michelle O’Hegarty, Daniel Owusu, Lindsey Page, Eric Pevzner, Mary Pomeroy, Ian W. Pray, Elizabeth M. Rabold, Jared R. Rispens, Phillip Salvatore, Cuc H. Tran, Jeni Vuong, Ashutosh Wadhwa, Sarah Willardson, Sherry Yin, Anna Yousaf implemented the study protocol, developed standards of practice, and collected data and specimens from the field.

Michelle Banks, Angela C. Dunn, Alicia Fry, Aron J. Hall, Hannah L. Kirking, Scott A. Nabity, Amy Schumacher, Jacqueline E. Tate, Ryan P. Westergaard, and Dongni Ye provided overall project coordination and leadership.

Hannah E. Reses, Mark Fajans, Garrett Fox, Allison Binder, and Victoria T. Chu curated the data.

Hannah E. Reses, Mark Fajans, Scott H. Lee, Charles M. Heilig, and Scott A. Nabity drafted the original manuscript.

## Utah

Davis County (UT) Health Department: Heather Gibb, Sara Hall

Summit County (UT) Health Department: Carolyn Rose

Salt Lake County (UT) Health Department: Dagmar Vitek, Ilene Risk, Lee Cherie Booth, Jeff Sanchez, Madison

Clawson, Tara Scribellito, Ha Khong, Carlene Claflin, Theresa Beesley, Victoria Castaneda

Utah Department of Health: Nathan LaCross, Robyn Atkinson-Dunn, COVID-19 response team

## Wisconsin

North Shore Health Department: Kala Hardy, Christine Cordova, Kevin Rorabeck, Kathleen Platt

City of Milwaukee Health Department: Catherine Bowman, Nancy Burns, Barbara Coyle, Elizabeth Durkes,

Carol Johnsen, Jill LeStarge, Erica Luna-Vargas, Sholonda Morris, Mary Jo Gerlach, Jill Paradowski, Bill Rice,

Michele Robinson, Virginia Thomas, Keara Jones, Chelsea Watry, Richard Weidensee, Jeanette Kowalik, Heather Paradis, Julie Katrichis

Wauwatosa Health Department: Laura Conklin, Paige Bernau, Emily Tianen

Wisconsin Department of Health Services: COVID-19 response team

City of Milwaukee Laboratory SARS-CoV-2 Testing Team: Jordan Hilleshiem, Beth Pfotenhauer, Manjeet Khubbar, Jennifer Lentz, Zorangel Amezquita-Montes, Kristin Schieble, Noah Leigh, Joshua Weiner, Tenysha Guzman, Kathy Windham, and Julie Plevak

## Centers for Disease Control and Prevention

CDC COVID-19 Response Team: Claire Midgley, Melissa Rolfes, Mayer Antoine, Adebowale Ojo, Fiona Havers Laboratory Task Force: Brandi Limbago, Michelle Owens, Wendi Kuhnert-Tallman, Jeff Johnson, Collette Leaumont, Gayle Langley

Field Investigations Team: Jenny Milucky, Emily Weston, Jessica Smith, Michelle Johnson Jones, Jennifer Huang, Erin Moritz, Aubrey Gilliland, Laura Calderwood, Jennifer Imaa, Kaytlin Renfro, Allison Miller, Katie Bantle, Margaret Williams, Stacy Thorne, Jana Manning, Micha Ghertner, Adriane Niare, Yoonjae Kang, Charlotte D. Kaboré

## Tables

**Supplemental Table 1.**
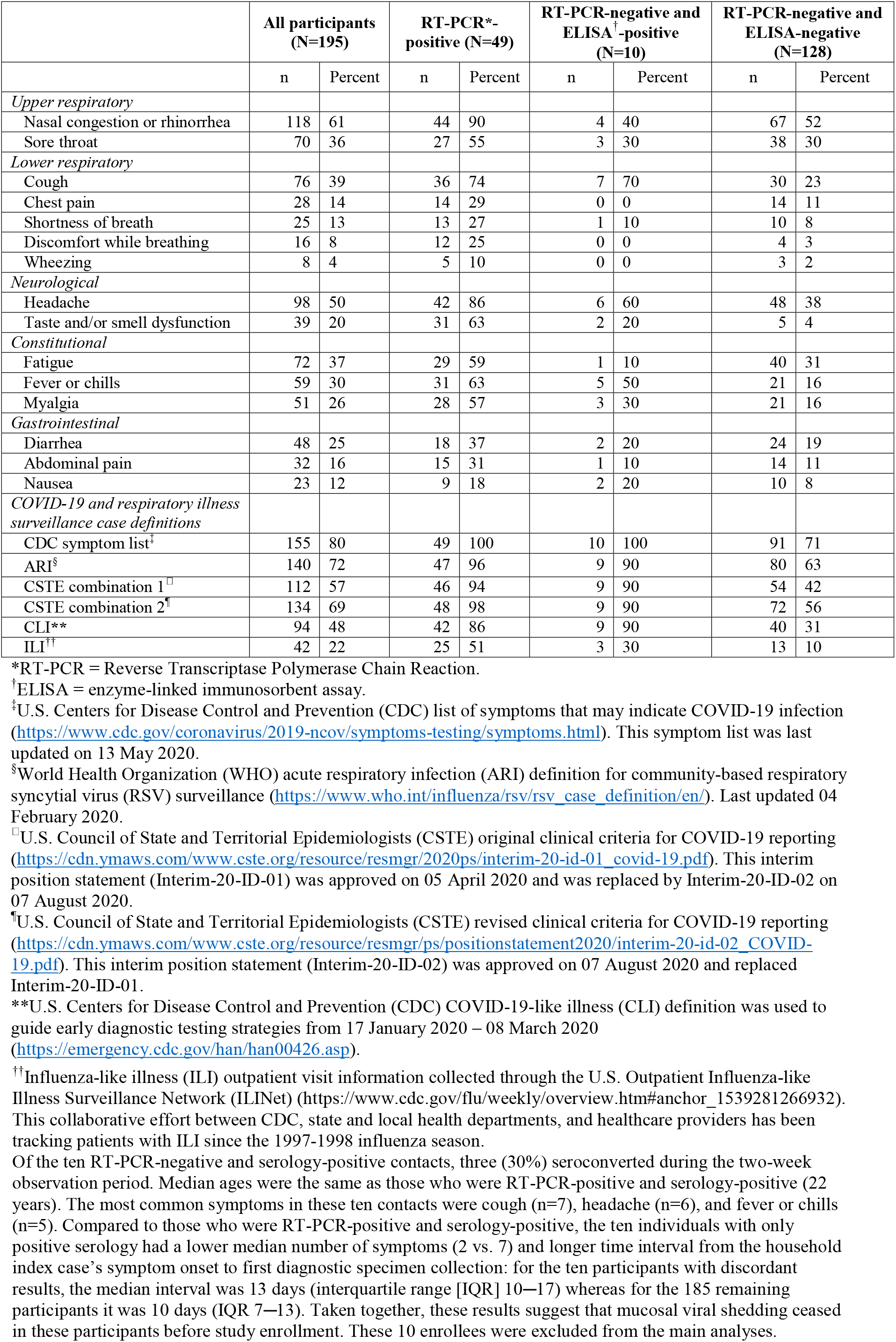
Individual COVID-19 symptoms and existing case definitions by 2019-nCoV Real-Time RT-PCR assay and a SARS-CoV-2 spike protein enzyme-linked immunosorbent assay (ELISA) results in Utah and Wisconsin, United States, March–May 2020

