## supplemental tables for "Performance of Existing and Novel Surveillance Case Definitions for COVID-19 in the Community"

**Supplemental Table 1. Individual COVID-19 symptoms and existing case definitions by 2019-nCoV Real-Time RT-PCR assay and a SARS-CoV-2 spike protein enzyme-linked immunosorbent assay (ELISA) results in Utah and Wisconsin, United States, March–May 2020**

|  | **All participants (N=195)** | | **RT-PCR*-positive (N=49)** | | **RT-PCR-negative and ELISA**^†^**-positive (N=10)** | | **RT-PCR-negative and ELISA-negative (N=128)** | |
| --- | --- | --- | --- | --- | --- | --- | --- | --- |
|  | n | Percent | n | Percent | n | Percent | n | Percent |
| *Upper respiratory* |  |  |  |  |  |  |  |  |
| Nasal congestion or rhinorrhea | 118 | 61 | 44 | 90 | 4 | 40 | 67 | 52 |
| Sore throat | 70 | 36 | 27 | 55 | 3 | 30 | 38 | 30 |
| *Lower respiratory* |  |  |  |  |  |  |  |  |
| Cough | 76 | 39 | 36 | 74 | 7 | 70 | 30 | 23 |
| Chest pain | 28 | 14 | 14 | 29 | 0 | 0 | 14 | 11 |
| Shortness of breath | 25 | 13 | 13 | 27 | 1 | 10 | 10 | 8 |
| Discomfort while breathing | 16 | 8 | 12 | 25 | 0 | 0 | 4 | 3 |
| Wheezing | 8 | 4 | 5 | 10 | 0 | 0 | 3 | 2 |
| *Neurological* |  |  |  |  |  |  |  |  |
| Headache | 98 | 50 | 42 | 86 | 6 | 60 | 48 | 38 |
| Taste and/or smell dysfunction | 39 | 20 | 31 | 63 | 2 | 20 | 5 | 4 |
| *Constitutional* |  |  |  |  |  |  |  |  |
| Fatigue | 72 | 37 | 29 | 59 | 1 | 10 | 40 | 31 |
| Fever or chills | 59 | 30 | 31 | 63 | 5 | 50 | 21 | 16 |
| Myalgia | 51 | 26 | 28 | 57 | 3 | 30 | 21 | 16 |
| *Gastrointestinal* |  |  |  |  |  |  |  |  |
| Diarrhea | 48 | 25 | 18 | 37 | 2 | 20 | 24 | 19 |
| Abdominal pain | 32 | 16 | 15 | 31 | 1 | 10 | 14 | 11 |
| Nausea | 23 | 12 | 9 | 18 | 2 | 20 | 10 | 8 |
| *COVID-19 and respiratory illness surveillance case definitions* |  |  |  |  |  |  |  |  |
| CDC symptom list^‡^ | 155 | 80 | 49 | 100 | 10 | 100 | 91 | 71 |
| ARI^§^ | 140 | 72 | 47 | 96 | 9 | 90 | 80 | 63 |
| CSTE combination 1^‖^ | 112 | 57 | 46 | 94 | 9 | 90 | 54 | 42 |
| CSTE combination 2^¶^ | 134 | 69 | 48 | 98 | 9 | 90 | 72 | 56 |
| CLI****** | 94 | 48 | 42 | 86 | 9 | 90 | 40 | 31 |
| ILI^††^ | 42 | 22 | 25 | 51 | 3 | 30 | 13 | 10 |

*RT-PCR = Reverse Transcriptase Polymerase Chain Reaction.

^†^ELISA = enzyme-linked immunosorbent assay.

^‡^U.S. Centers for Disease Control and Prevention (CDC) list of symptoms that may indicate COVID-19 infection (<https://www.cdc.gov/coronavirus/2019-ncov/symptoms-testing/symptoms.html>). This symptom list was last updated on 13 May 2020.

^§^World Health Organization (WHO) acute respiratory infection (ARI) definition for community-based respiratory syncytial virus (RSV) surveillance (<https://www.who.int/influenza/rsv/rsv_case_definition/en/>). Last updated 04 February 2020.

^‖^U.S. Council of State and Territorial Epidemiologists (CSTE) original clinical criteria for COVID-19 reporting (<https://cdn.ymaws.com/www.cste.org/resource/resmgr/2020ps/interim-20-id-01_covid-19.pdf>). This interim position statement (Interim-20-ID-01) was approved on 05 April 2020 and was replaced by Interim-20-ID-02 on 07 August 2020.

^¶^U.S. Council of State and Territorial Epidemiologists (CSTE) revised clinical criteria for COVID-19 reporting (<https://cdn.ymaws.com/www.cste.org/resource/resmgr/ps/positionstatement2020/interim-20-id-02_COVID-19.pdf>). This interim position statement (Interim-20-ID-02) was approved on 07 August 2020 and replaced Interim-20-ID-01.

**U.S. Centers for Disease Control and Prevention (CDC) COVID-19-like illness (CLI) definition was used to guide early diagnostic testing strategies from 17 January 2020 – 08 March 2020 (<https://emergency.cdc.gov/han/han00426.asp>).

^††^Influenza-like illness (ILI) outpatient visit information collected through the U.S. Outpatient Influenza-like Illness Surveillance Network (ILINet) (https://www.cdc.gov/flu/weekly/overview.htm#anchor_1539281266932). This collaborative effort between CDC, state and local health departments, and healthcare providers has been tracking patients with ILI since the 1997-1998 influenza season.

Of the ten RT-PCR-negative and serology-positive contacts, three (30%) seroconverted during the two-week observation period. Median ages were the same as those who were RT-PCR-positive and serology-positive (22 years). The most common symptoms in these ten contacts were cough (n=7), headache (n=6), and fever or chills (n=5). Compared to those who were RT-PCR-positive and serology-positive, the ten individuals with only positive serology had a lower median number of symptoms (2 vs. 7) and longer time interval from the household index case’s symptom onset to first diagnostic specimen collection: for the ten participants with discordant results, the median interval was 13 days (interquartile range [IQR] 10─17) whereas for the 185 remaining participants it was 10 days (IQR 7─13). Taken together, these results suggest that mucosal viral shedding ceased in these participants before study enrollment. These 10 enrollees were excluded from the main analyses.
